# Single unit-derived connectivity networks in tuberous sclerosis complex reveal propensity for network hypersynchrony driven by tuber-tuber interactions

**DOI:** 10.1101/2024.05.09.24306995

**Authors:** Aswin Chari, Amanda E Hernan, J Matthew Mahoney, Rachel Thornton, M Zubair Tahir, Martin M Tisdall, Rod C Scott

## Abstract

Network hypersynchrony is emerging as an important system-level mechanism underlying seizures, as well as cognitive and behavioural impairments, in children with structural brain abnormalities. We investigated patterns of single neuron action potential behaviour in 206 neurons recorded from tubers, transmantle tails of tubers and normal looking cortex in 3 children with tuberous sclerosis. The patterns of neuronal firing, on a neuron-by-neuron (autocorrelation) basis did not reveal any differences as a function of anatomy. However, at the level of functional networks (cross-correlation), there is a much larger propensity towards hypersynchrony of tuber-tuber neurons that in neurons from any other anatomical site. This suggests that tubers are the primary drivers of adverse outcomes in children with tuberous sclerosis.

## Introduction

Tuberous sclerosis complex (TSC) is a common genetic disorder, usually caused by loss of function germline mutations in *TSC1* or *TSC2* genes.^1^ The phenotype includes a range of neurological symptoms including seizures, cognitive impairments, and behavioural disorders that are associated with cortical brain tubers.^1^ These tubers share genetic mutations with and are histologically identical to focal cortical dysplasia type 2b and consist of a tuber core and transmantle tail in the subjacent white matter that often extends towards the ventricular surface.^2^ The seizures, cognitive impairments, and behavioural disorders in TSC are attributed to tubers, although the mechanisms by which tubers disrupt function remains uncertain, particularly as cells in both the tuber and normal appearing cortex will have the germline mutation.^1^ We suggest that abnormal structural architecture in the tuber underlies abnormal neuronal network dynamics that in turn mediate the emergent phenotypes, including seizures and the cognitive and behavioural dysfunction associated with TSC.

Epilepsy is characterised by spontaneous, unprovoked seizures.^3^ Seizures are characterised by abnormal synchronous firing of cortical areas. Previous studies evaluating action potential firing or calcium dynamics in cultured induced pluripotent stem cells from patients with TSC have suggested that there is network hypersynchrony measured as increased functional connectivity between neurons.^4,5^ However, these studies are unable to address whether the level of hypersynchrony differs as a function of histological structure in patients. There is therefore a gap in our understanding of the basic mechanisms of synchronisation between what happens at the level of individual neurons in patient brains and the abnormal dynamics supporting hypersynchrony at the level of large volumes of brain tissue. Understanding these mechanisms may influence downstream clinical decision-making including approaches to reduce seizures (eg resective epilepsy surgery) and strategies to ameliorate the other co-morbidities associated with TSC.

The processes of synchronisation and de-synchronisation of correlated brain activity are fundamental to normal brain functions.^6^ Excessive synchronization characterising seizures has been hypothesised to derive from abnormal network properties that constrain the dynamics of the epileptic brain to become paroxysmally pathologically hypersynchronous.^7^ However, there is growing evidence from mesoscale dynamics, such as intracranial EEG, that neural activity outside of seizures is also abnormal, and specifically implicate a network poised to become synchronous.^8,9^ Because the brain is not hypersynchronous in these inter-ictal periods, we must infer the propensity to generate hypersynchronous activity from non-hypersynchronous epochs. The ability of correlated brain activity to generate hypersynchronous activity is a property we term *synchrogenicity*.

To examine the relationship between functional connectivity and *synchrogenicity*, we took advantage of recent progress in microelectrode technology, that has facilitated the simultaneous recording of clinically relevant local field potential (LFP) and single- and multi-unit activity during stereoelectroencephalography (SEEG), which is often required in TSC prior to the formulation of a strategy for resective epilepsy surgery.^10^ We studied single unit electrophysiological activity in three children with TSC undergoing evaluation for epilepsy surgery to examine whether neurons in the *in vivo* network, outside of the time of seizures, have a propensity for hypersynchrony and, if so, whether neurons in specific areas (the tuber core, the tuber tail or radiologically unaffected cortex) drive abnormal network dynamics.

We hypothesised that functional firing patterns analysed on a neuron-by-neuron basis would be similar across histological regions given that TSC is a germline disorder, likely affecting the biophysical properties of each neuron equally, but that correlated firing in tuber cores would be markedly abnormal when compared to other regions due to the disrupted network structure. We studied functional firing patterns using a statistical model for neural spike trains,^11,12^ which we used to define two novel, network-based, interictal measures of synchrogenicity: ***autonomous synchrogenicity*** and ***network synchrogenicity*** (Figure 1). These properties define how the single-unit network is expected to respond to synchronous inputs, by either propagating them or suppressing them as a function of either neuron-autonomous or network-based functional firing characteristics, respectively. We study the synchrogenicity of histologically distinct brain regions within and around cortical tubers, which are strongly correlated with abnormal firing and are thought to be a major driver of hypersynchrony. We show that network synchrogenicity is driven by interaction between neurons within and between tubers, demonstrating that these histologically disordered regions have a high level of influence on the synchronization of neuronal networks. This supports the view that both seizures and the other neurological symptoms in TSC are driven by abnormal neural dynamics that arise from disrupted structural architecture within tubers.^13^

**Figure 1:**
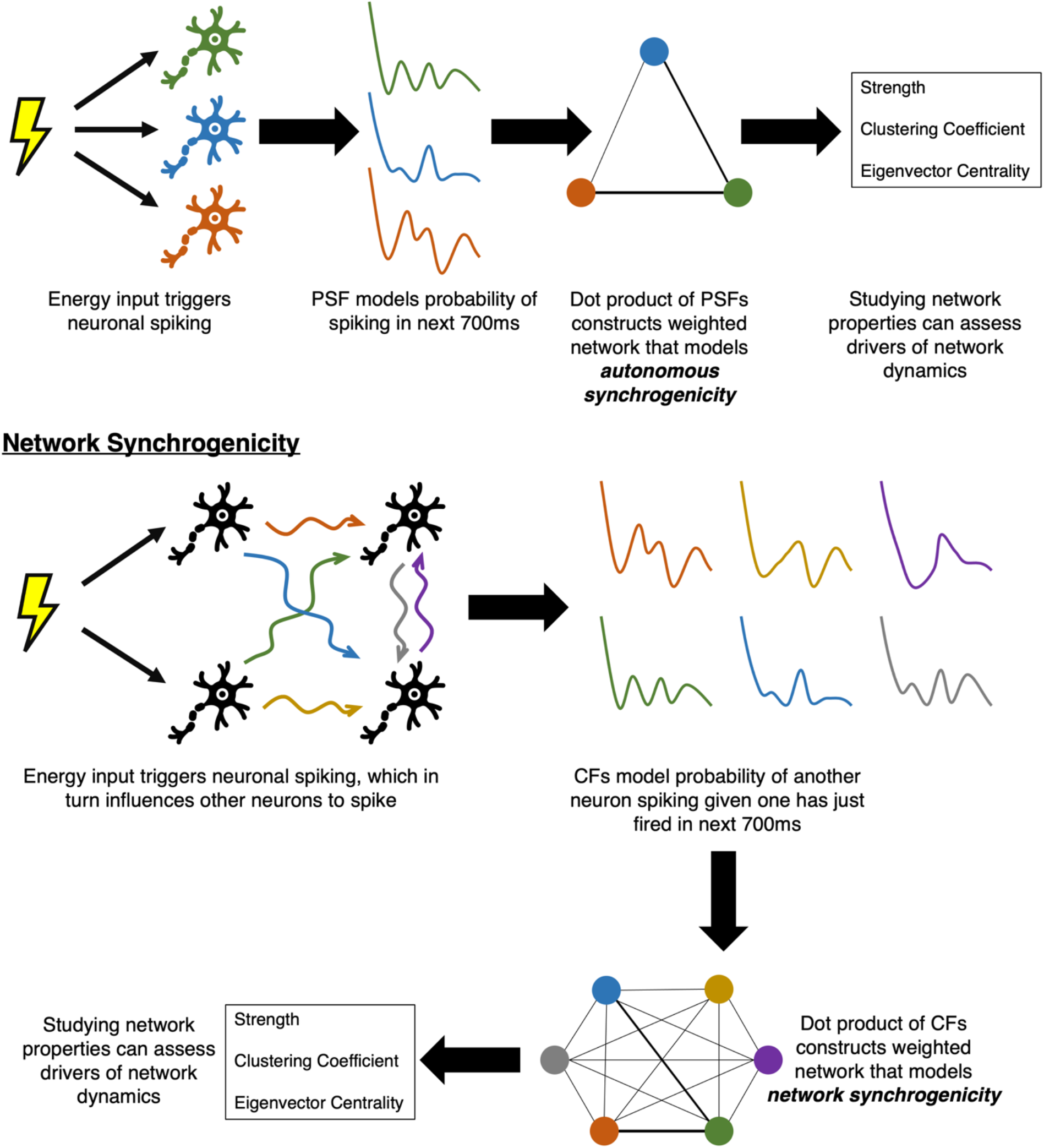
Schematic of analytic techniques for assessment of autonomous and network synchrogenicity (propensity to generate hypersynchronous activity) from single unit spike trains. It involves the modelling of single unit spike trains to generate post-spike filters (PSF) and coupling filters (CFs). The correlations of these can assess autonomous and network synchrogenicity respectively and building networks from these can assess drivers of synchrogenicity.

## Methods

This prospective study was approved by the London-Brent Research Ethics Committee on behalf of the UK Health Research Authority (IRAS ID 255823). In line with UK law, the parents of all included patients provided informed consent for study participation.

### Participants

Three patients with TSC were included in the study (Table 1). All were selected to undergo stereoelectroencephalography (SEEG) as part of their clinical work-up for drug resistant epilepsy. Clinical decision making, including decision to undergo SEEG and the number and location of electrodes were not affected by involvement in the study. All underwent predominantly frontal explorations with some electrodes extending into the insula and parietal cortices.

**Table 1:**
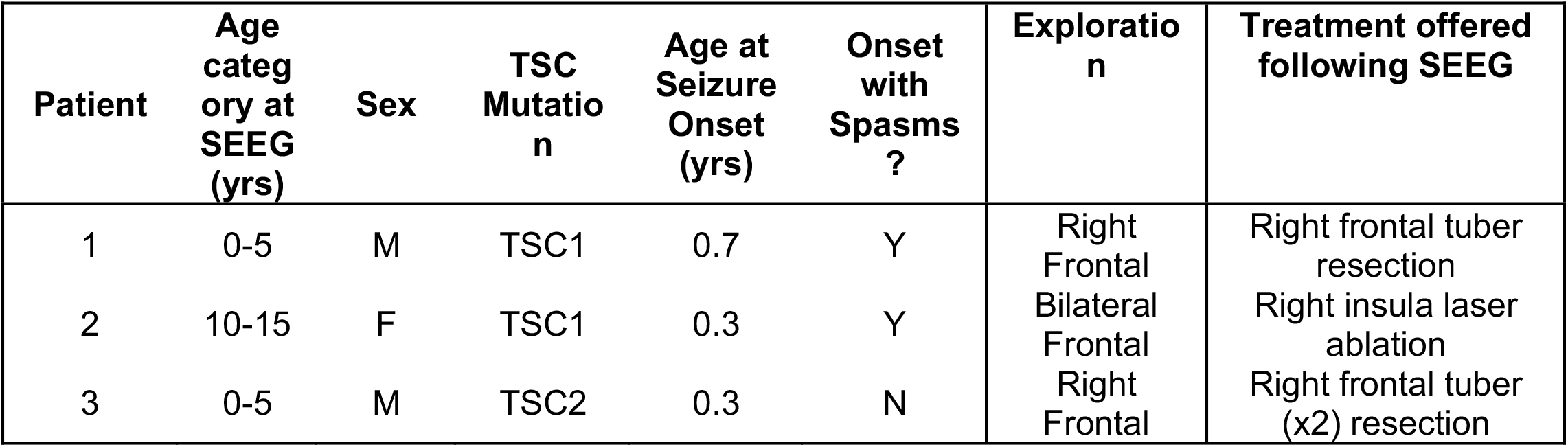
Demographic details of the 3 included patients.

| Patient | Age category at SEEG (yrs) | Sex | TSC Mutation | Age at Seizure Onset (yrs) | Onset with Spasms ? | Exploration | Treatment offered following SEEG |
| --- | --- | --- | --- | --- | --- | --- | --- |
| 1 | 0-5 | M | TSC1 | 0.7 | Y | Right Frontal | Right frontal tuber resection |
| 2 | 10-15 | F | TSC1 | 0.3 | Y | Bilateral Frontal | Right insula laser ablation |
| 3 | 0-5 | M | TSC2 | 0.3 | N | Right Frontal | Right frontal tuber (x2) resection |

### Microelectrode Recording and Data Processing

Enrolled patients underwent insertion of three (NNAIS05), two (NNAIS06) and 5 (NNAIS07) hybrid micro-macro electrodes (AdTech MM16A-SP05X-000), each with 10 microelectrode contacts along the shaft (Figure 2a). Hybrid electrodes replaced existing planned macroelectrode trajectories and were chosen based on the length of the planned electrode trajectories; the hybrid micro-macro electrodes had a fixed active length of 28mm and it was therefore not possible to specifically target the purported epileptogenic tubers. Post hoc, it was revealed that none of the isolated neurons were from putative epileptogenic tubers that were subsequently resected or ablated. Macro channels were connected to the clinical system (Natus Quantum) to record at 2kHz whilst micro channels were connected to a research system with pre-amplification (Ripple Summit) to record at 30kHz. The same white matter macro contacts were chosen as ground and reference for both systems and connected to each other to reduce noise.

**Figure 2:**
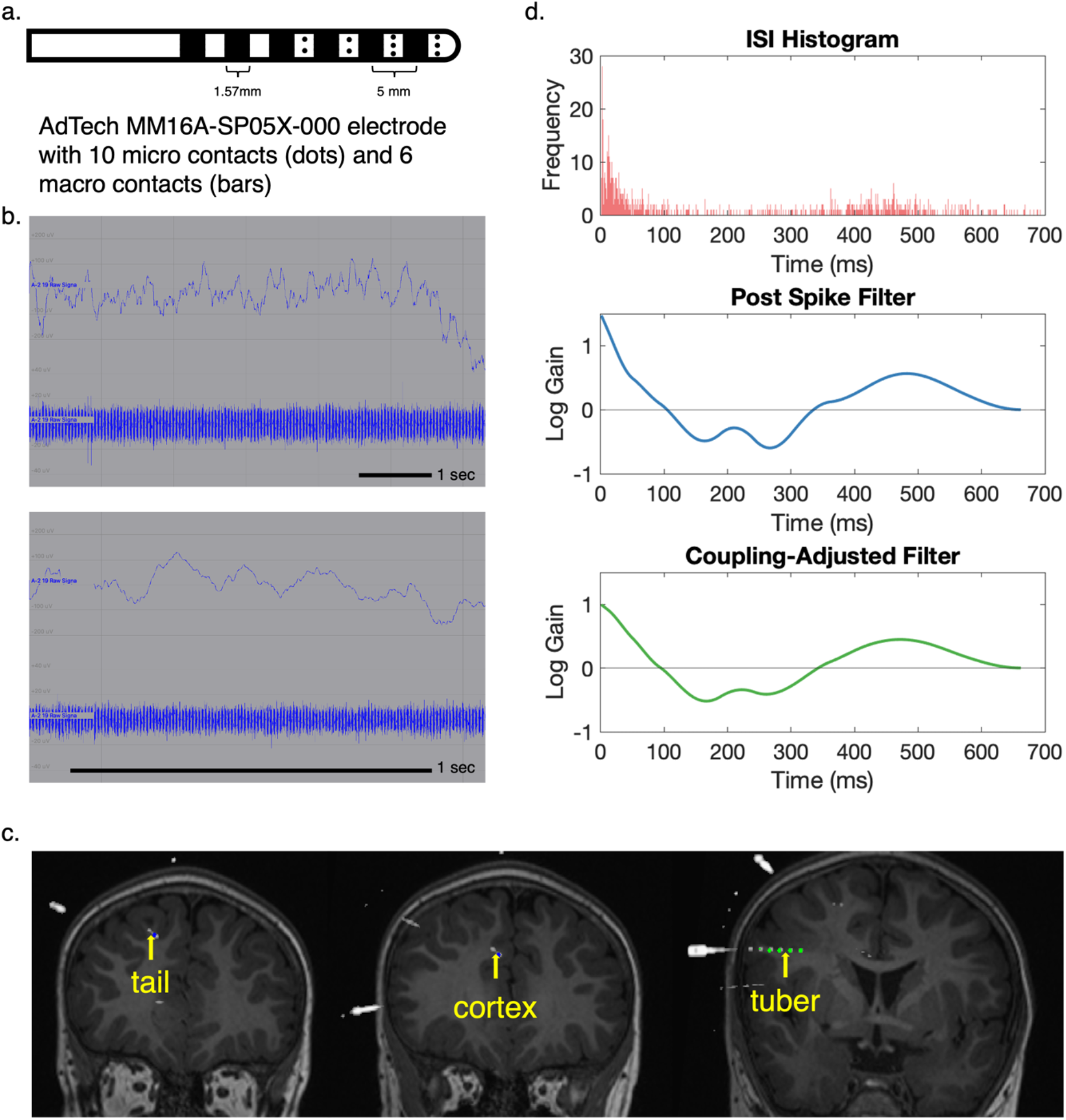
Summary of methods. (a) Schematic of AdTech hybrid micro-macro SEEG electrodes used in this study. (b) Raw traces of recordings from single microelectrode channel showing LFP (1-100 Hz, upper) and high frequency (500+ Hz, lower) traces. The top panel shows a 10s montage whilst the lower panel shows a 1s montage. (c) Fused coronal T1-weighted MRI & post-operative CT scan in one patient showing examples of contact locations labelled as cortex, tuber and tail. The microelectrode contacts (yellow arrow) are adjacent to the macroelectrode contacts (green/blue dots). (d) Example of an inter-spike interval (ISI) histogram, post spike filter and coupling-adjusted post spike filter from a neuron on the same channel shown in b.

Microelectrode recordings were conducted for up to 30 mins per day for each recording day possible. This was in the awake state during the day whilst the child was sitting in bed, either watching the television or playing puzzle/activity games. Recordings were performed before the first recorded seizure in all patients.

Microelectrode recordings (Figure 2b) were processed using Matlab R2020b, FieldTrip & WaveClus3.^14,15^ On channels with acceptable impedances on impedance testing (50-400 kOhms) and local field potential (LFP) visible at low frequencies, a discrete fourier transform filter (at 50,100 and 150Hz) was applied to reduce line noise. Spikes were extracted using positive and negative detection at a 4 standard deviation above the noise threshold and subsequently sorted using WaveClus3 and manually selected based on a combination of the temperature plots and firing rate. Putative units were excluded if >2% of the inter spike intervals (ISIs) were <2ms and their firing rates were less than 0.5Hz. As we wanted to sample all interictal connectivity, interictal epileptiform discharges (IEDs) were not sought out and continuous data was used.^16^

Each spike train was assigned a location based on the fusion of the post-operative CT to MRI as either within the tuber, in the transmantle tail or in radiologically unaffected cortex (Figure 2c). This was possible as the predominant SEEG implantation strategy at our institute is to sample putative tubers, the depths reaching the white matter tails, with occasional electrodes in normal areas of cortex guided by the non-invasive presurgical evaluation data. Spike properties were assessed using the frequency, the spike half width and coefficient of variance of the ISIs (standard deviation of ISIs / mean ISI).^17^

Generalised linear models were used to generate post-spike filters (PSFs) and coupling filters (CFs) for each neuron within each recording session (Figure 1). These filters are statistically robust models of auto- and cross-correlation properties of the spike trains, allowing an assessment of the statistical influence of past firing (PSF) and the past firing of other neurons (CF) on future firing.^11,12,18^ The GLM has a baseline firing rate parameter controlling the average propensity of a neuron to fire and all other effects are encoded as a time-varying gain functions called filters that multiplicatively modulate the firing rate. The PSF encodes the modulation of a neuron’s firing rate given it has just fired, whilst the CF encodes the modulation of a neuron’s firing rate given that another neuron has just fired (Figure 1). The CF therefore acts as a directed connectivity measure between the influencing neuron and target neuron. Following normalisation of the filters to enable shape comparisons, two main analyses were performed as follows.

First, to quantify variation in auto- or cross-correlation structures defined by the PSF and CFs, we decomposed the filter time series using principal component analysis (PCA). Differences in the first 2 principal components were compared between histological regions (3 groups for PSFs and 9 groups for CFs).

Second, we developed novel, network-based measures to assess *autonomous synchrogenicity* and *network synchrogenicity* (Figure 1). Because healthy brains must constantly switch between synchronous and asynchronous states, we hypothesised that the mechanisms of hypersynchrony are due to abnormal responses to synchronous inputs. When a region of brain tissue receives inputs that drive neurons to fire at the same time, the dynamical response to those inputs will produce outputs that can either be synchronous or asynchronous. During interictal, asynchronous activity, we cannot directly observe the drive toward synchrony, but we can estimate this response using the fitted GLMs from above.

#### Autonomous synchrogenicity

Given two neurons with normalised PSFs, *f*_*i*_(*t*) and *f*_*j*_(*t*), we define the dot product of their PSFs as

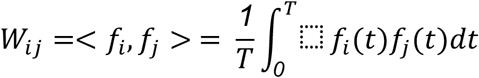

where T is duration of the PSFs. *W*_*ij*_ measures the similarity of the predicted future firing of neurons i and j given that they fired simultaneously. For *n* neurons, this *n* x *n* matrix of pairwise dot products *W* = [*W*_*ij*_]_⬚_can be viewed as a weighted adjacency matrix for an abstract network among neurons from which we computed the following three common network measures using the Brain Connectivity Toolbox^19^:

- Strength (*i*.*e*., weighted degree): Node strength is the sum of weights of edges connected to the node. High strength implies a densely connected node.
- Eigenvector Centrality: Eigenvector centrality is a self-referential measure of centrality; nodes have high eigenvector centrality if they connect to other nodes that have high eigenvector centrality. A high eigenvector centrality implies a high level of influence on the network.
- Clustering Coefficient: The clustering coefficient is the weight of triangles around a node and is equivalent to the weight of node’s neighbours that are neighbours of each other. A high clustering coefficient implies that a node’s neighbours are all densely connected to each other, while a low clustering coefficient indicates that a node’s neighbours are spread out across the network.

Each abstract network measure separately encapsulates the idea that some nodes are more tightly interconnected within the abstract network. Functionally, neurons with high values for strength and eigenvector centrality and low values for clustering coefficient are predicted by the GLM to produce time-correlated outputs with many other neurons in the ensemble, provided their inputs cause them to spike at the same time. This could happen, for example, if a subset of neurons had highly stereotyped oscillatory PSFs. Thus, these network measures capture our notion of *autonomous synchrogenicity* and comparing between histological regions facilitates an assessment of which regions drive the *synchrogenicity* of the network.

#### Network synchrogenicity

Given two pairs of neurons with normalised CFs, *f*_*ij*_(*t*) and *f*_*kl*_(*t*), we define the dot product of their CFs as

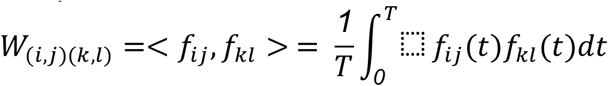

where T is duration of the CFs. *W*_(*ij*)(*k*)_ measures the similarity of predicted future firing of neurons j and k given that neurons i and l fired simultaneously. The complete matrix of pairwise dot products *W* = [*W*_(*ij*)(*k*)_]_⬚_can be viewed as a weighted adjacency matrix for an abstract network. Note that the neurons i, j, k, and l need not be distinct.

For *n* neurons, this *n*^*2*^ x *n*^*2*^ matrix of pairwise dot products *W* = [*W*_(*ij*)(*k*)_]_⬚_can be viewed as a weighted adjacency matrix for an abstract network among pairs of neurons, from which we can compute strength, clustering coefficient, and eigenvector centrality, as above. Analogous to *autonomous synchrogencity* above, neuron pairs with high values for strength and eigenvector centrality and low values for clustering coefficient are predicted by the GLM to produce time-correlated outputs with many other neurons in the ensemble, provided their inputs cause them to spike at the same time, this time based on their cross-correlation patterns. The important distinction in this case is that, unlike *autonomous synchrogenicity*, the inputs and outputs can be different neurons, so synchronous inputs in one part of the network can propagate to generate synchronous outputs at another part of the network. Thus, these network measures capture our notion of *network synchrogenicity*. Again, comparing between histological regions facilitates an assessment of which regions drive the *synchrogenicity* of the network.

To compare network measures between the different histological regions, we used standard linear models. To adjust for networks of different sizes, we compared the ranks of the network measures within each subject and each recording session, normalised to a scale between 0 and 1. Statistical analyses were performed using parametric tests. P-values <0.05 were considered statistically significant.

## Code and Data Availability

Code used for this study is available at https://github.com/aswinchari/SingleUnitConnectivity/ and data (the single unit spike trains after spike sorting, subsequent filters and networks) are available at https://osf.io/ab39m/.

## Results

In total, 206 units were isolated across 7 recording sessions (total 3h and 5 mins) in the 3 patients, all performed within the first three days (day 0, day 1 and day 2) of SEEG implantation. Of these, 79 were in normal cortex, 53 were in the body of a tuber and 74 were in the tail of the tuber.

### Isolated neurons show largely typical features of pyramidal cells with only subtle behavioural differences between the groups

Across all units, the neurons showed features of typical pyramidal cells with mean firing rate of 1.7Hz (SD 1.8) and mean spike half width of 0.18ms (SD 0.02) (Figure 3a). There was no separate cluster of smaller spike half width and higher frequency cells to indicate interneurons. There were no differences in spike frequency (one-way ANOVA, f(2,203)=0.01, p=0.99) and spike half width (one-way ANOVA, f(2,203)=1.86, p=0.16) by location Figure 3b). The coefficient of variation of ISIs was different (one-way ANOVA, f(2,203)=6.64, p=0.002), with pairwise comparison using Tukey’s method showing that the coefficient of variation was higher in the normal cortex compared to both the tuber (p=0.003) and tail (p=0.01). This suggests a more stereotyped pattern of firing of the tuber & tail neurons compared to normal cortical neurons.

**Figure 3:**
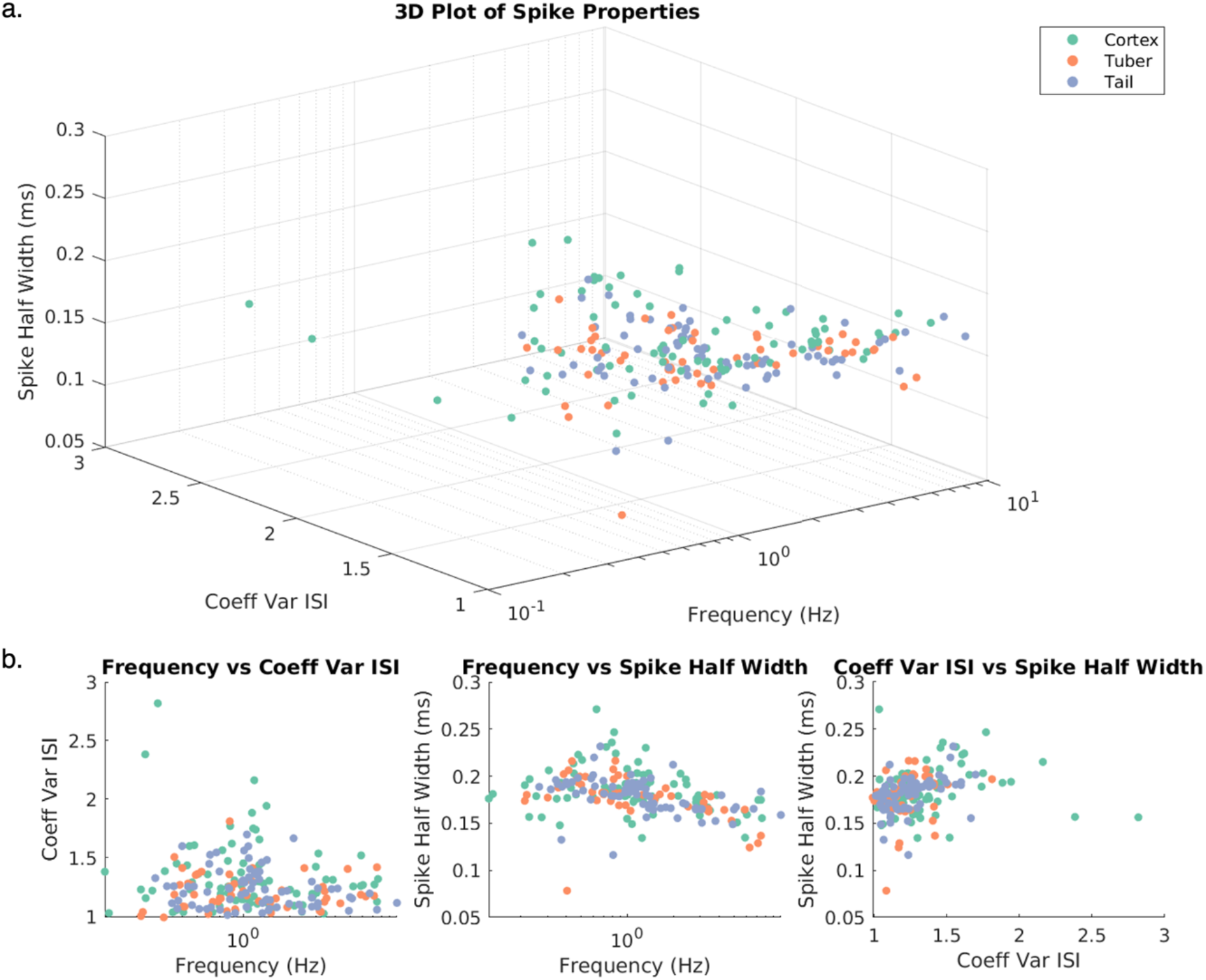
Spike properties of isolated single units. (a) 3D plot of three common spike properties used to sort between cell types, coded by location of the cells. (b) 2D plots of each combination of properties. Both illustrate that (i) there are no clear clusters of different cell types to indicate pyramidal cells and interneurons and (ii) there are no differences in spike frequency and half-width between cell types, but the cortical spikes have a higher coefficient of variation of ISIs compared to spikes in the cortex and tail.

### Post spike filter properties indicate similar neuron autocorrelation structure across regions

To ascertain whether there were differences in firing patterns of individual neurons in the different locations, the post-spike filters (PSFs) for each of the 206 neurons were decomposed using principal component analysis (PCA), of which the first 2 principal components explained 63% of variability in the data (Figure 4a). The first principal component describes a bursting neuron with bursts occurring approximately every 120ms. The second principal component describes initial bursting preceding a refractory period lasting about 150ms followed by no further autocorrelation structure. The scores for PC1 and PC2 are a measure of similarity between an individual neuron’s PSF shape and the canonical shapes shown (Figure 4c). There were no differences between neurons in different histological areas, indicating that neurons in the tuber core, tail and radiologically unaffected cortex all have similar auto-correlation structure (Figure 4b).

**Figure 4:**
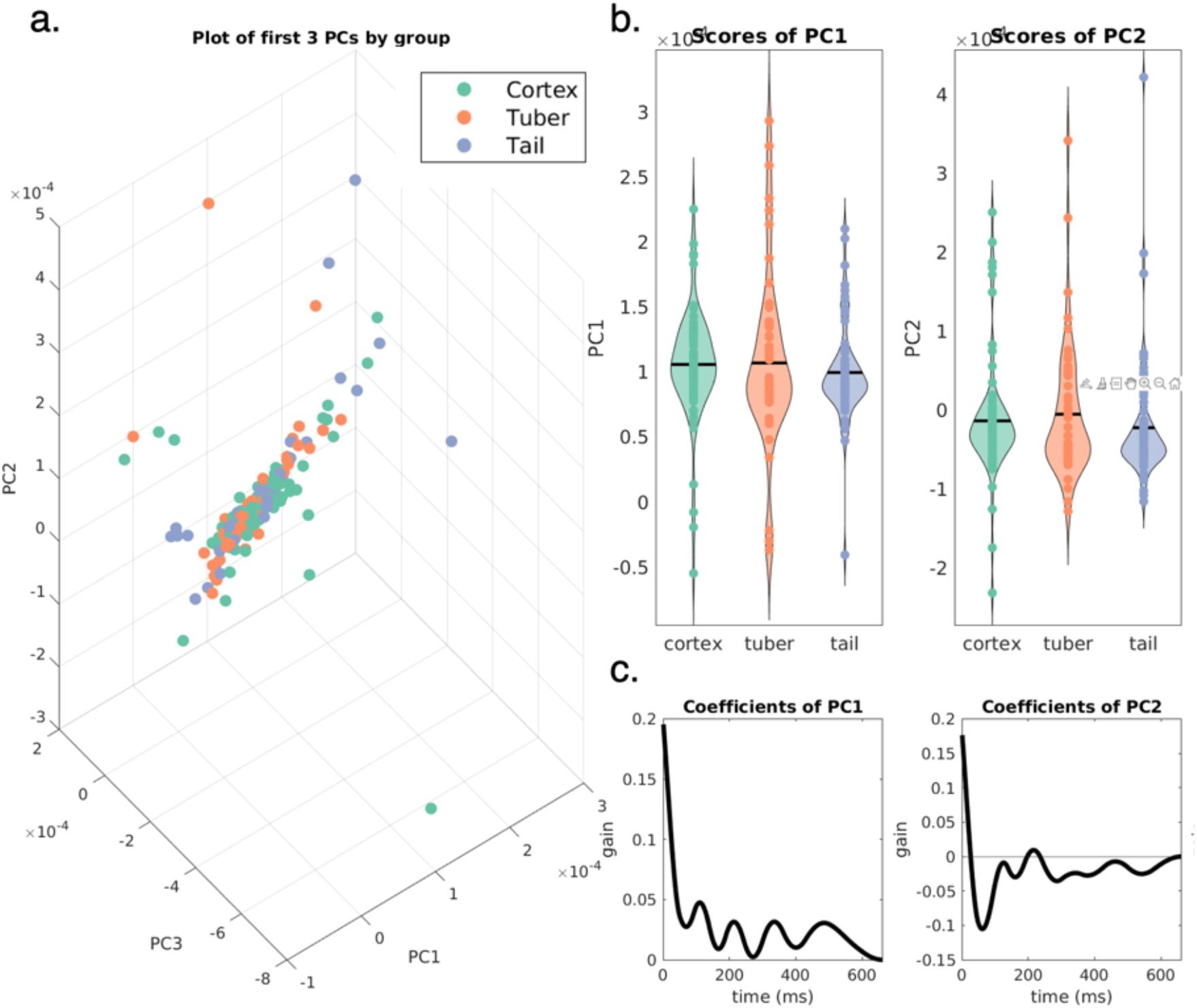
Principal component analysis of post spike filters. PCA was performed to decompose PSFs from 206 neurons. (a) 3D plot of the first 3 PC scores, coded by location of the cells. (b) Scores of the first 2 PCs (which explained 63% of the variance) by location of the cells. There were no significant differences in the PC scores across groups. Black bars indicate means. (c) Coefficients of these first 2 PCs indicating the nature of the modulation conferred by each PC.

### Coupling filter properties indicate dynamic non-stereotyped interactions between neurons which are not different between groups

Next, we built coupling filters (CFs) to quantify neuronal firing co-modulation between pairs of simultaneously recorded neurons in our dataset and similarly decomposed the CFs of the 9302 pairs of neurons using PCA, of which the first 2 principal components explained 25% of the variability in the data (Figure 5). The CF describes cross-correlation structure after autocorrelation structure has been subtracted. Therefore, this provides different information to correlation between PSFs and is a more direct measure of how neurons influence each other. PC1 describes an initial burst of firing followed by modulation of firing at alpha frequencies (increased propensity to fire 200 and 350 ms after the initial action potential) (Figure 5c). The scores for PC1 (one-way ANOVA, f(8,9293)=2.84, p=0.004) were statistically significantly different between groups (Figure 5b). Pairwise comparisons using Tukey’s method showing lower PC1 score in ‘tuber-cortex’ filters compared to ‘tuber-tuber’ (p=0.03) and ‘cortex-cortex’ (p=0.006), indicating lower alpha modulation in tuber-cortex coupling filters compared to the 2 other types of filters. There were no differences between PC2 scores as a function of the histological region (Figure 5b).

**Figure 5:**
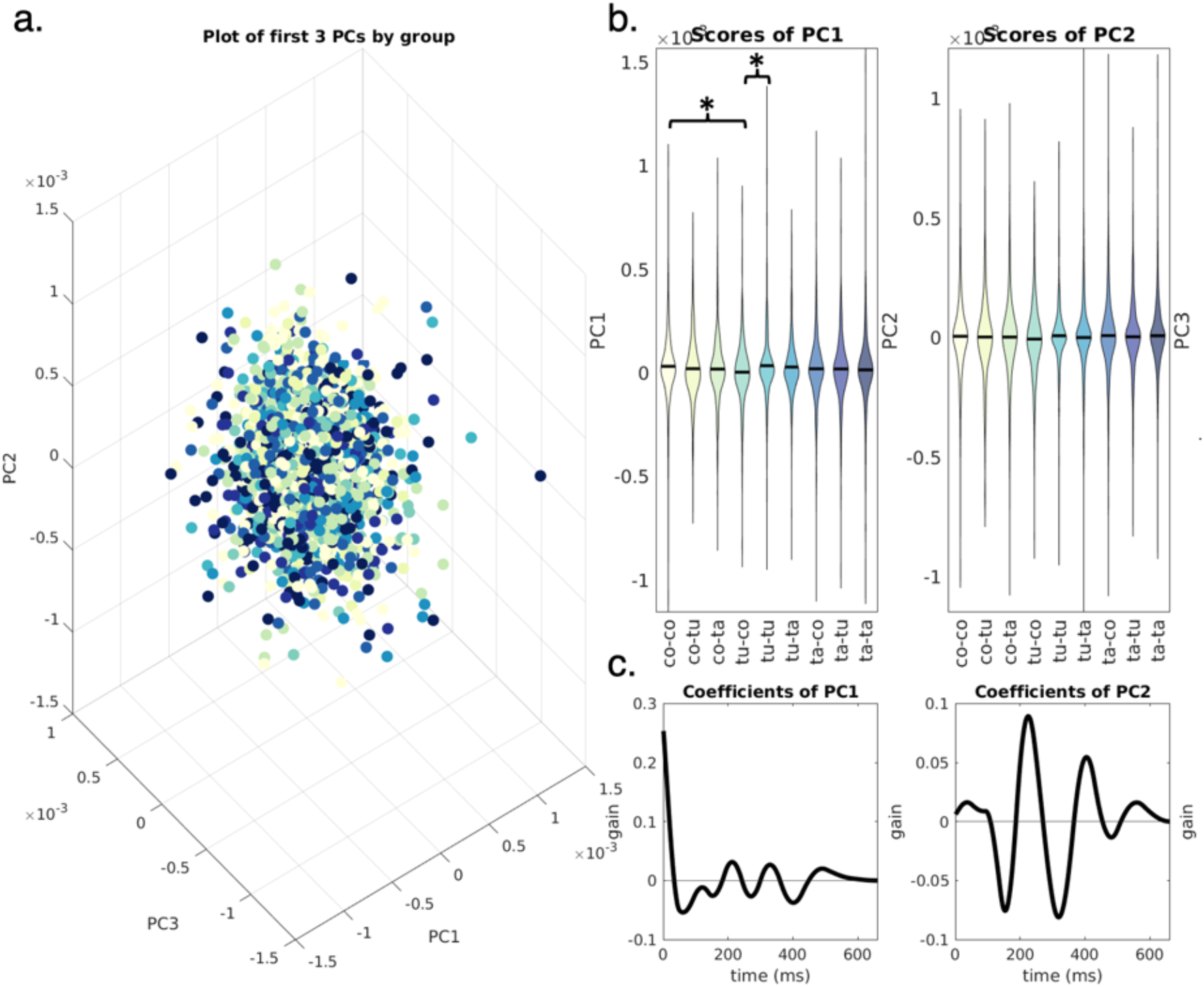
Principal component analysis of coupling filters. PCA was performed to decompose CFs from 9301 pairs of neurons. (a) 3D plot of the first 3 PC scores, coded by nature of the CFs. (b) Scores of the first 2 PCs (which explained 25% of the variance) by nature of the CFs. Black bars indicate means. There were significant differences (*) between PC1 scores between the denoted groups. (c) Coefficients of these first 2 PCs indicating the nature of the modulation conferred by each PC.

Overall, the low variance explained by the first 2 PCs indicate a wide range of ways in which pairs of neurons influence each other, reflecting the dynamic nature of these interactions that facilitate diverse functions. The fact that there are only subtle differences in the PC1 scores indicates that there are no specific stereotyped cross-correlation phenotypes that seem to drive network dysfunction in TSC.

### Hypersynchrony may be driven by high network synchrogenicity of tuber-tuber interactions

The previous analyses evaluated single neuron and pairwise firing behaviours in isolation, showing, at most, modest differences in firing properties during interictal brain activity between groups. However, because of prior findings showing subtle functional network differences in interictal recordings at other spatial scales^8,9^, we extended our approach to evaluate the expected dynamical response to synchronous inputs.

Specifically, we constructed two types of abstract networks. The first was among PSFs encoding synchronous output firing due to synchronous input spikes, which we termed *autonomous synchrogenicity*. The second was among CFs encoding the propagation of synchronous activity due to synchronous inputs, which we termed *network synchrogenicity*. We analysed network properties of nodes within these networks and assessed whether abstract network properties differed between groups. Both network types and the rationale for these analyses are explained in more detail in the methods.

Networks constructed from PSFs in individual recording sessions revealed no difference in 3 network measures, namely strength (one-way ANOVA, f(2,203)=1.75, p=0.18), clustering coefficient (one-way ANOVA, f(2,203)=0.17, p=0.84), and eigenvector centrality (one-way ANOVA, f(2,203)=1.76, p=0.18) between units that were in tuber, tail and cortex. This indicates no inherent differences in *autonomous synchrogenicity* between neurons in the tuber, tail and cortex.

However, when similar networks were constructed for CFs, there were significant differences across all 3 graph metrics; strength (one-way ANOVA, f(8,9293)=27.5, p=1×10^-42^), clustering coefficient (one-way ANOVA, f(8,9293)=7.1, p=2×10^-9^) and eigenvector centrality (one-way ANOVA, f(8,9293)=16.8, p=5×10^-25^) (Figure 6a-c, left panel). Pairwise comparisons using Tukey’s method revealed multiple differences (Figure 6a-c, right panel). The most robust changes across the groups were increased strength (p<3×10^-4^ compared to all other groups), decreased clustering coefficient (p<5×10^-3^ compared to all other groups) and increased eigenvector centrality (p<9×10^-5^ compared to all other groups) for the tuber-tuber CFs, indicating increased *network synchrogenicity* being driven by tuber-tuber interactions.

**Figure 6:**
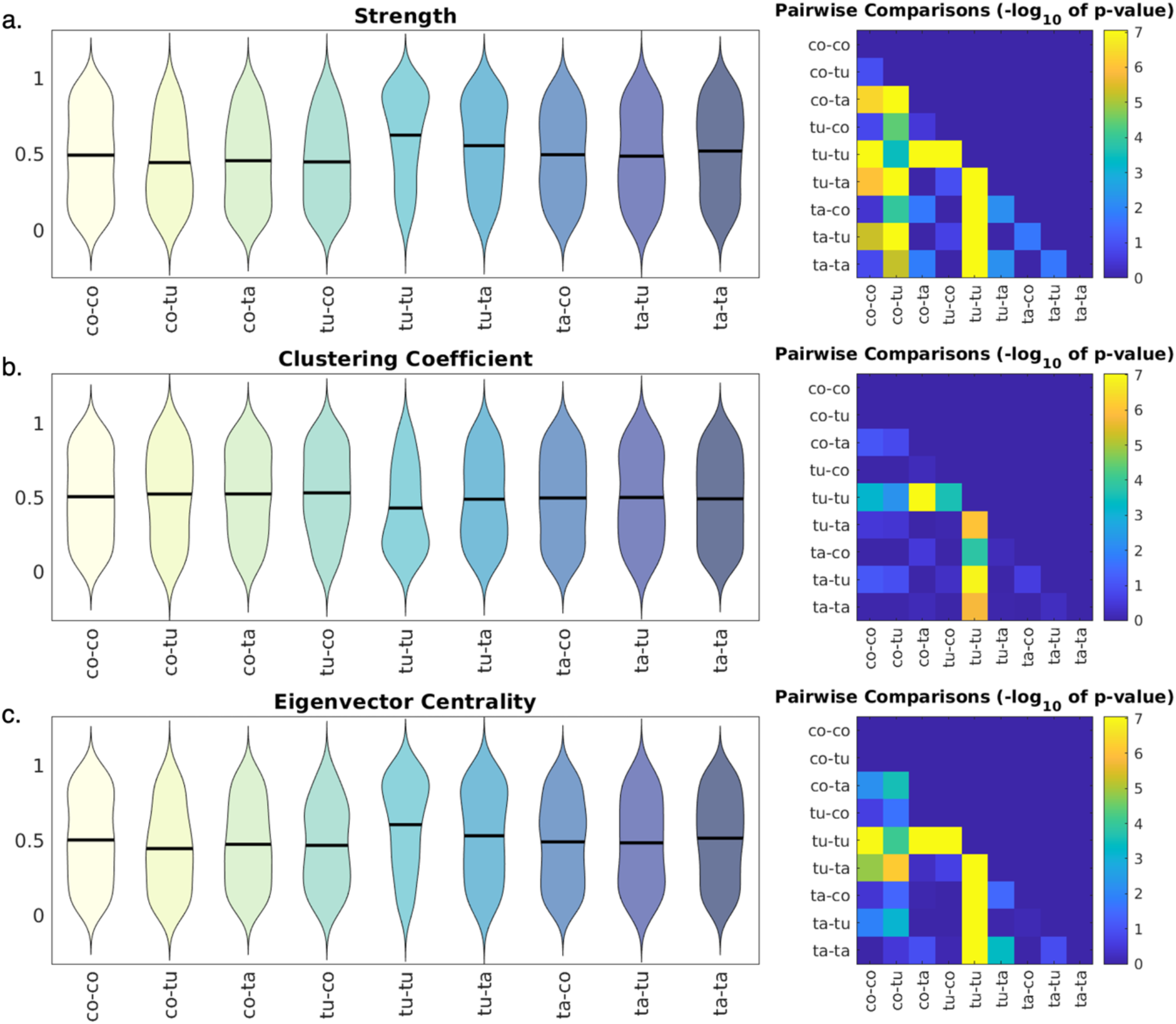
Properties of the networks derived from coupling filters differ by type of edge. Node (a) strength, (b) clustering coefficient and (c) eigenvector centrality vary by type of coupling filter in eFC networks across sessions. Black bars indicate means. The panels on the right show pairwise comparisons, indicating that the most striking features are increased strength, decreased clustering coefficient and increased eigenvector centrality of the tuber-tuber filters compared to all other groups. (co = cortex, tu = tuber, ta = tail).

## Discussion

In this single-unit electrophysiology study in 3 children with TSC, we reveal that individual neuronal firing properties, as modelled through the shapes of their statistically robust post spike and coupling filters, are similar in different histological regions. Using novel network analyses in interictal periods, we show that the *autonomous synchrogenicity* of these neurons is also not different between regions but, when assessing *network synchrogenicity*, we identify that the propensity for network hypersynchrony, which may underlie the seizures and associated comorbidities, is driven by tuber-tuber interactions in abnormally connected functional networks.

### Post-spike and coupling filters shapes are not different between regions

There are at least two potential explanations for why we fail to see differences in the shapes of the post-spike and coupling filters between regions when deconstructed using principal component analysis. The first is that all neurons have the mutation, similar biophysical properties and thus respond similarly to inputs. This is consistent with cell culture and in-vitro experiments in animal models of TSC that neurons have abnormal shape and electrophysiological properties even when there is no overt histological malformation.^4,5^ Therefore, the similarity in autocorrelated firing patterns may not be surprising as the germline mutation in TSC likely affects all neurons, even those outside tubers. The second is that the brain regions being sampled simply do not have differences in firing patterns similar to the lack of difference between normal and abnormally structured cortical firing.^18^

The first two principal components explain only a small proportion of the variance in the filters (63% for PSFs and 25% for CFs), underscoring the importance of varied, non-stereotyped responses of neurons to inputs that facilitate the emergence of varied and adaptable phenotypes which can be both functional (e.g. cognitive processes) and dysfunctional (e.g. seizures).

### Abstract networks reveal differences in network synchrogenicity but not autonomous synchrogenicity

When we constructed networks that model the responses to simultaneous inputs, we identified no differences in network properties between histological regions when assessing *autonomous synchrogenicity*. However, we showed differences in network properties of networks constructed from CFs, indicating that *network synchrogenicity* was driven by tuber-tuber interactions. These findings indicate that the propensity to hypersynchrony is not driven by a specific group of neurons in the tuber, tail or cortex but instead by a complex functional organization wherein the co-firing of neurons throughout the ensemble is more similar to the co-firing of pairs of neurons within or between tubers.

Action potential firing dynamics are fundamental to brain function, and it is widely accepted that phenotypes are mediated through neural network activities.^13^ Therefore, evaluating how neural dynamics are altered in disease can lead to improved understanding of disease pathogenesis and subsequently to development of novel therapies. There is a large literature on action potential dynamics in animal models confirming that dynamical patterns are abnormal across a range of insults and that the nature of the alterations can predict phenotypes.^13,18,20^ However, the translation of these techniques to humans has been limited and to our knowledge this is the first time that these approaches have been evaluated in children. Furthermore, rodent models of TSC lack the histological features of TSC, namely tubers, making questions about connectivity within tubers and between tuber and adjacent tissue impossible to address pre-clinically.^21^ Indeed, many studies have explored TSC firing abnormalities using iPSCs from patients, but these studies also cannot address tuber-specific alterations in network properties.

Importantly, this study uses only TSC patient data and therefore, we cannot infer that the network is inherently prone to hypersynchrony compared to a healthy brain but that, within the observed single-unit functional network, the tuber-tuber interactions drive the propensity for hypersynchrony through their network functional connectivity rather than inherent differences in firing properties of individual neurons.

### Implications of tuber-tuber interactions driving hypersynchrony

The network dynamics identified in this study have implications in a number of domains. Firstly, in terms of identifying tubers and networks responsible for seizures in TSC, there may be a complex interplay between multiple tubers driving hypersynchrony and therefore, highlight the need to rethink the paradigm of identifying specific epileptogenic tubers. Indeed, clinical evidence suggests that seizures often arise from multiple tubers and require more complex resection strategies.^22,23^ Further evidence may be gained from linking tuber connectivity metrics to epileptogenicity and associating these with post-resection outcomes.

The findings from this network approach are unique and build upon work from other studies in TSC. Using SEEG, Alexander et al found increased inter- and intra-tuberal connectivity in the beta and gamma bands during seizures.^22^ Similarly, Yu et al found increased proportion of positive cortico-cortical evoked potentials (CCEPs) between tubers compared to tuber-cortex and cortex-tuber, implying enhanced connectivity between cortical tubers.^24^ They further linked the strength of these connections to the epileptogencity of the tubers, with more positive CCEPs between epileptogenic and early propagation tubers. They also found that the CCEP positivity rate was highest in the centre of the tubers, suggesting a gradient of epileptogenicity within tubers, with a peak in the cores. This concept of the tuber core driving the dynamics has also been shown in a dynamic causal modelling study, which showed that ictal and interictal dynamics were best supported by a model where an impulse was triggered at the tuber core.^25^ This is supported by another single patient single unit study which showed a gradient of epileptogenicity within tubers, with more neurons being modulated by interictal epileptiform discharges in the tuber compared to the periphery.^26^ Together, these works support a complex interaction within and between tubers that needs to be considered before deciding on treatment strategies including surgical resection of potentially epileptogenic tubers.

These findings may also be of interest in non-TS contexts. If other epileptogenic lesions such as focal cortical dysplasia have similar pathophysiological mechanisms and EEG signatures,^27–29^ it may allow better localisation of epileptogenic lesions using interictal data. This is particularly important as a recent study has shown that current neurophysiological definitions of the seizure onset zone may be deficient as the proportion of these contacts resected during SEEG-guided resection does not correlate with outcome; in that study, post-operative identification of pathological tissue was the only predictive factor.^30^

### Does microelectrode data have a role in the future of invasive presurgical evaluation?

The findings of this and other recent studies showcase the technical ability to extract single unit data from microelectrodes embedded into clinically used SEEG electrodes.^10,26,31^ They provide a proof of principle that such recordings may hold promise for understanding epileptic and malformed cortex. As outlined in an earlier review, there are a number of ways such recordings could help, including improving the localisation of the epileptogenic zone, allowing localisation using interictal recordings and providing a substrate for closed loop neuromodulation technologies.^10^

Current barriers to such clinical translation include technical difficulty, processing power and a currently rudimentary understanding of the interpretation of the data. In terms of technical difficulty, this study (and other similar ones^32,33^) have highlighted difficulties in optimising recording conditions and equipment for single unit recordings from human brains. Improvements in technology, both at the electrode and recording amplifier stages will facilitate robust recordings that are reliable and have good signal-to-noise ratios. This study requires hours of post-recording processing to generate the networks and improvements in computational power will aid almost real-time processing of such metrics, that may then facilitate their clinical utility. Lastly, more work in this area will elucidate clinically important biomarkers of the epileptogenic zone (or other metrics) that can then be used to guide surgical management and/or neuromodulation to optimise patient outcomes.

## Conclusion

In this study of 3 patients with TSC, we show using microelectrode recordings, robust filters and functional connectivity networks from single unit spike trains that there is a propensity to hypersynchrony across the TSC brain, driven not by abnormal neuronal firing but by functional interactions of neuronal firing within and between tubers within the functional connectivity network. This provides a proof of principle that microelectrode recordings can be used to construct networks and analysis of these networks may be helpful in increasing our understanding of the pathophysiological mechanisms in TSC and related disorders and ultimately could be useful to guide clinical management.

## Data Availability

https://github.com/aswinchari/SingleUnitConnectivity/

